# Prefrontal Cortex Definitions and Their Use in Distinguishing Pornography Addicted Juveniles

**DOI:** 10.1101/2021.06.22.21259291

**Authors:** Pukovisa Prawiroharjo, Rizki Edmi Edison, Hainah Ellydar, Peter Pratama, Sitti Evangeline Imelda Suaidy, Nya’ Zata Amani, Diavitri Carissima, Ghina Faradisa Hatta

## Abstract

**Background and aims:** Increasing popularity of Internet has exposed our children pornography addiction. As in other types of addiction, it affects a brain region known as prefrontal cortex (PFC), which is important in executive functions and inhibitory control. However, this region was loosely defined, and there was no consensus for that definition. We aimed to use volumetric MRI in finding the defining region of PFC which would be suitable in distinguishing pornography addicted juveniles.

**Methods:** We enrolled 30 juveniles (12-16 y.o.) consisting of 15 pornography addiction and 15 non-addiction subjects. We proposed several models of PFC definition from mix-and-matched subregions, consisting of orbitofrontal (OFC), inferior frontal gyrus (IFG; pars orbitalis, opercularis, and triangularis), dorsolateral PFC (DLPFC), and anterior cingulate (ACC). Suitable PFC definition was defined as models which volume statistically different between both groups. Brain volumetric was measured using 3D-T1 3T MRI images and analyzed using FreeSurfer® for automatic cortical reconstruction and brain segmentation (recon-all command).

**Results:** We found significant differences between groups in 6 models, which mainly included OFC, ACC, and DLPFC, with models devoid of DLPFC had lowest mean differences.

**Conclusion:** The most suitable definition of PFC for pornography addiction study should consist of OFC, ACC, and especially DLPFC. Inferior frontal gyrus pars orbitalis was not necessary for this purpose, but may increase effect size if it is included.

## Introduction

Increasing popularity of Internet has exposed our children to various contents unsuitable for them. A survey in 2011–2012 by Indonesian Ministry of Communication and Informatics showed that more than 50% respondents (10-19 y.o.) had been exposed to pornography, with 14% of them accessing pornographic websites voluntarily. Another survey in 2016 showed that ±97% elementary students of 4–6^th^ grade in Jakarta and surrounding area had been exposed to pornography.[1] Addiction to pornography has been found on individuals as young as 10 and as old as 22, but most of them are juveniles aged 13–18 years. Hilton and Watts, based on various neuroscientific studies of addictions, postulated that pornography addiction may cause not only chemical, but also anatomical and pathological changes, in the brain. Collectively called as hypofrontal syndromes, they signify disruption of “braking system” and may include compulsivity, impulsivity, emotional lability, and impaired judgment.[2] Similarly, American Society of Addiction Medicine, although not specifically mentioned pornography, treats behavioral addiction similar as substance addiction, in that both affects neurotransmission, hippocampal-cortical circuit interaction, brain reward structure, and triggers addictive behaviors.[3] Pornography addiction is also known as cause of marriage problems, social isolation, career loss,[4] and sexual crimes.[5,6]

Substance addiction are known to involve a region of brain called prefrontal cortex (PFC), which is important in executive functions and exerts inhibitory control on behavior (“braking system”).[7–9] Not surprisingly, this finding was consistent in pornography addiction despite there are far fewer neuroscientific studies regarding it.[10–12]

Numerous processes take place in PFC, contributing to neuropsychological functions such as emotion, cognitive, and behavior. Interruption in these processes might lead individuals to lose their self-control and salience, such in the case of addiction [9]. The highly integrated processes in PFC subregions themselves, together with other brain regions, makes it hard to tell specific function of individual subregion. As suspected, studies about injecting cocaine intravenously to cocaine-addicted individuals resulted in activation of numerous PFC subregions.[13,14]

However, currently the term “prefrontal cortex” is somewhat loosely defined. Fuster defined it simply as “cortex of the anterior pole of the mammalian brain”.[15] Some studies only mentioned three subregions of PFC (out of five circuits[16–18]) which are involved in addiction, i.e. dorsolateral prefrontal cortex (DLPFC), orbitofrontal cortex (OFC), and anterior cingulate gyrus (ACC).[7–9] Some other studies did not define it in details, which means it is hard to do precise systematic review or metaanalysis on this matter due to possible bias in definition of PFC. This might be somewhat mitigated in studies of substance addiction due to its sheer number of publications, but for pornography addiction, which has scarce evidences, this inconsistency may become a potential problem.

This study aimed to use volumetric MRI in finding the defining region of PFC which would be suitable in distinguishing pornography addicted juveniles, as attempt to standardize neuroscientific studies of pornography addiction in the future.

## Materials and Methods

### Ethical consideration

The study was approved by Health Research Ethical Committee of Faculty of Medicine Universitas Indonesia (Clearance No. 1155/UN2.F1/ETIK/2017) and conducted in accordance to Helsinki Declaration. No subject was confronted with pornographic material in this study. Informed consent was obtained from all participants, represented by respective parents.

### Participants

As part of our previous study, we recruited 30 juveniles aged 12-16 years old during December 2017-February 2018, in various events held by YKBH in Bekasi, Indonesia. Using Pornography Addiction Test, a battery of neuropsychological test designed and validated by YKBH (see below),[19] we grouped the subjects into pornography addiction and non-addiction groups. Exclusion criteria were left-handed, verbal or language disorder, history of brain-related disorder or disease, head trauma, trauma during pregnancy or birth, developmental, psychological or neurological disorder, or mental illness.

### Pornography addiction screening

Pornography Addiction Test is a self-reported questionnaire designed by expert psychologist of YKBH to use in evaluating pornography addiction on Indonesian juveniles. Briefly, the questionnaire consisted of 99 questions composed from various studies of pornography addiction, evaluating four dimensions: time, motivation to use pornography, problematic pornography use, and consequences of addiction. Pornography addiction was defined as weighted score of greater than or equal to 32.[19]

### Definition of PFC

Previous studies mentioned five primary circuits of PFC: motor (originating in supplementary motor area), oculomotor (originating in frontal eye fields), dorsolateral prefrontal (DLPFC), orbitofrontal (OFC), and anterior cingulate (ACC). The former two are involved in motor activities, while the latter three are involved in executive and behavioral activities[16–18] and thus were often associated with addiction.[8] Therefore, we would focus to these regions in this study.

We proposed several models of PFC definition consisting of various mix-and-matched subregions, detailed in **Table 1**. We also included two subregions currently arguable whether to or not to be considered as PFC regions: Broca’s area and ACC. Broca’s area is anatomically located in inferior frontal gyrus (IFG), which is part of PFC, and a study indicated that IFG as a whole may have role in addiction, especially in self-control and attention.[9] However Broca’s area has been recognized for its exclusive role in speech.[20] On the other hand, ACC was classically considered as part of limbic cortex (not frontal lobe), but is commonly referred to when discussing PFC. Goldstein and Volkow (2011) defined ACC as part of prefrontal cortex,[9] while Weinstein and Lejoyeux treated it separatedly.[21] Classic definition of PFC would be model number C. All other models were mix-matched using the classic definition as pivot: model A-B pairs added Broca’s area, E-F pairs excluded IFG pars orbitalis, G-H pairs excluded OFC, and I-J pairs excluded DLPFC. All even models (B, D, F, H, and J) included ACC while odd models did not.

**Table 1.**
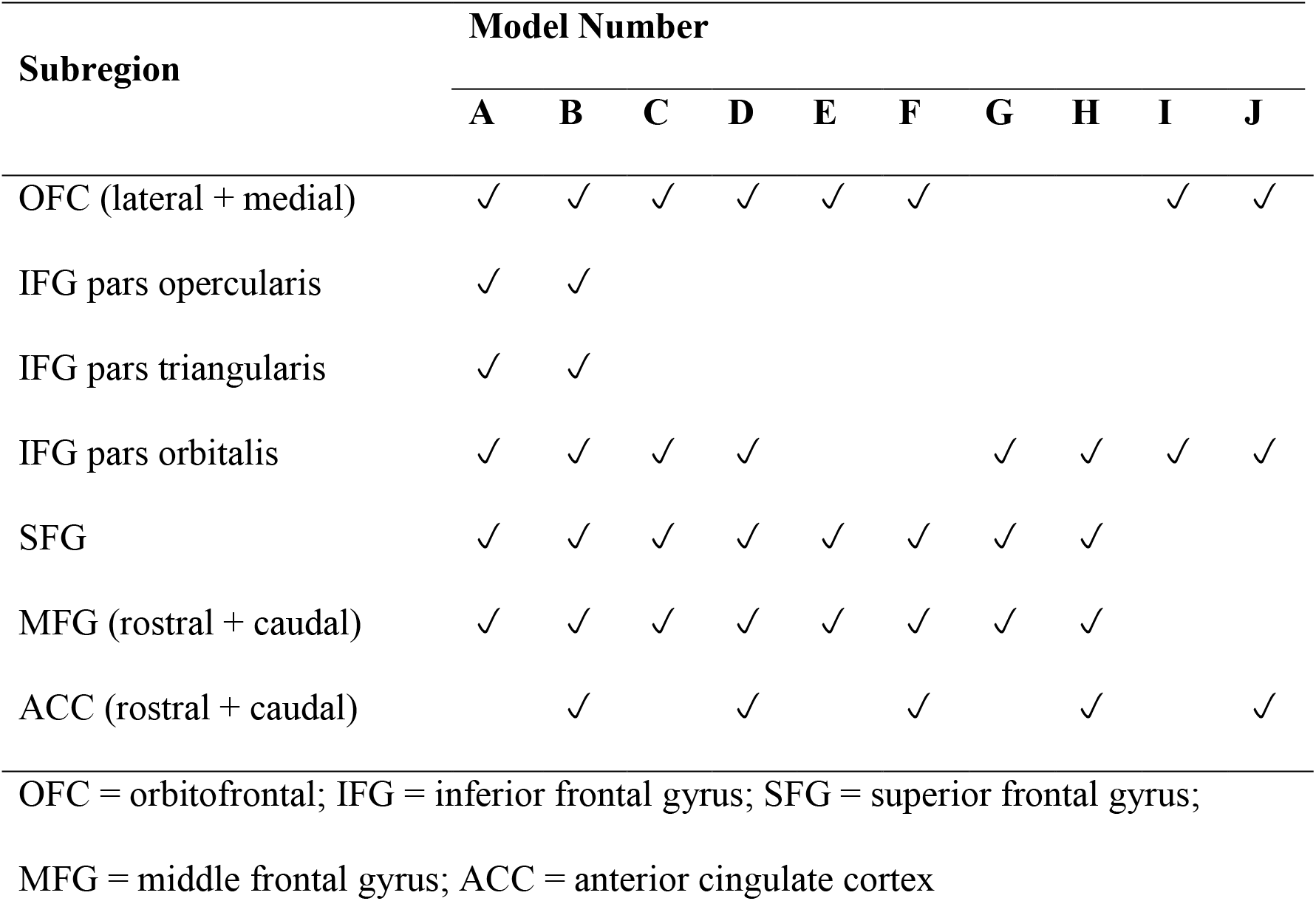
Investigated mixed-and-matched subregions (A-J) models defining prefrontal cortex

Due to limitation of FreeSurfer® labelling, we obtained volume of some regions from mathematical addition: OFC from its lateral and medial parts, MFG from its rostral and caudal parts, and ACC from its caudal and rostral parts. Additionally, Broca’s area was calculated from IFG pars opercularis and triangularis, while DLPFC was calculated from SFG and MFG.[22]

### Definition of “suitable” PFC definition

To find the suitable PFC definition for pornography addiction, we measured the volume of each our model in both groups, then normalized them into Brain Parenchymal Fraction (BPF, model volume divided by total intracranial volume).[23] However, as the regions we investigated would result in too small BPF, we had to use modified version of BPF by multiplying them with 1,000, resulting in what we dubbed as Brain Parenchymal Fraction per mill (BPF‰). We then compared the results between both groups. Significant statistical difference signified that corresponding model can be used to distinguish pornography addicts from non-addicts.

### Brain imaging acquisition and processes

To find volume of various brain subregions, we used MRI scans with GE-DISCOVERY® MR750 3 Tesla MRI scanner (8-channel-coil). We performed 3D T1-weighted sequence contiguous slice with the following parameters: repetition time = 8.3 sec; echo time = 3.2 sec; bandwidth = 31.3 kHz; field of view 24 × 24 mm; slice thickness = 1 mm; no gap (0); matrix 256 × 256 (frequency x phase), NEX 1; flip angle = 12º; time = 256 seconds. Images were then analyzed further using FreeSurfer® image analysis suite (http://surfer.nmr.mgh.harvard.edu/) for automatic cortical reconstruction and brain segmentation. FreeSurfer®’s N3 correction were also used to correct tissue signal inhomogeneity, to improve accuracy and robustness during cortical segmentation.[24] We obtained brain volume using the cross-sectional mode of recon-all flag -3T script of FreeSurfer version 6.0.[24] The script was used to improve diagnostic separability, accuracy, and reliability of cortical thickness by altering the parameters of FreeSurfer®’s internal N3 bias field correction.[24,25]

### Statistical analysis

We used Mann-Whitney tests for comparison between groups. Statistical significance was assumed on p < 0.05. All statistical analysis was performed using R environment version 3.4.4 on Microsoft® Windows® 10.

## Results

### Demographic Data

We enrolled 30 subjects (15 non-addiction vs 15 addiction group). Mean age was 13.27 ± 1.03 vs 13.80 ± 1.26 y.o. Both groups had matched age (p = 0.23).

### BPF in Subregions

Standing for DLPFC, only MFG showed significant lower BPF% in addiction group (p = 0.040, MD = -2.51). Calculated from SFG and MFG, DLPFC standalone did not result significant BPF% difference in addiction group (p = 0.071), although it also showed rather lower BPF%, marked by largest negative mean difference (MD = - 3,59) (**Table 2**).

**Table 2.** Region definition of investigated prefrontal cortex models.

| Subregion | BPF% |  | MD | p |
| --- | --- | --- | --- | --- |
|  | Non-addiction | Addiction |  |  |
|  | (n=15) | (n=15) |  |  |
| OFC (lateral + medial) | 20.93 ± 1.54 | 20.54 ± 1.22 | -0.39 | 0.468 |
| IFG Broca | 12.66 ± 1.53 | 12.56 ± 1.18 | -0.10 | 0.852 |
| IFG Total | 16.80 ± 1.81 | 16.55 ± 1.38 | -0.25 | 0.756 |
| SFG | 34.99 ± 1.99 | 33.91 ± 2.83 | -1.08 | 0.290 |
| MFG (rostral + caudal) | 36.15 ± 2.44 | 33.64 ± 2.96 | -2.51 | 0.040* |
| ACC (rostral + caudal) | 6.24 ± 0.77 | 5.80 ± 0.51 | -0.44 | 0.120 |
| DLPFC | 71.14 ± 3.73 | 67.55 ± 4.52 | -3.59 | 0.071 |

\* Statistically significant ( $p < 0.05$ )
OFC = orbitofrontal; IFG = inferior frontal gyrus; SFG = superior frontal gyrus;
MFG = middle frontal gyrus; ACC = anterior cingulate cortex

### BPF in Each Models

From the suggested models, six of them were potential to be used in juvenile pornography addiction: model B (p = 0.044), model C (p = 0.029), model D (p = 0.024), model E (p = 0.040), model F (p = 0.033), and model H (p = 0.040) (**Table 3**). In all models, BPF‰ of addiction group was significantly lower, marked by the negative mean differences. Largest mean difference was that of model B (MD = -4.57), followed closely by model D (MD = -4.57). Model I and J had smallest mean differences. (**Fig 1**).

**Table 3.**
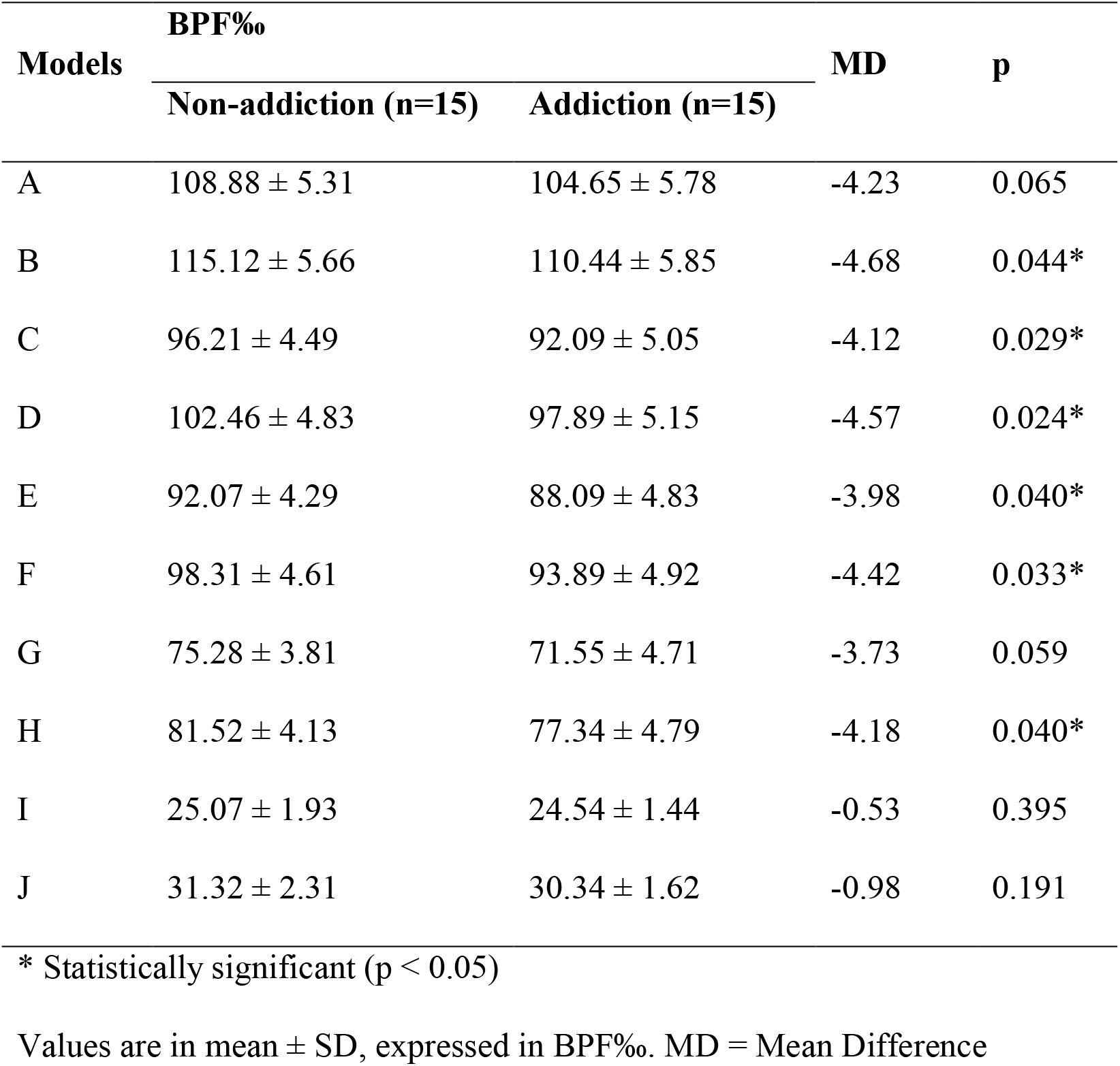
PFC model comparison between juvenile pornography addiction and non-addiction group.

**Fig 1.**
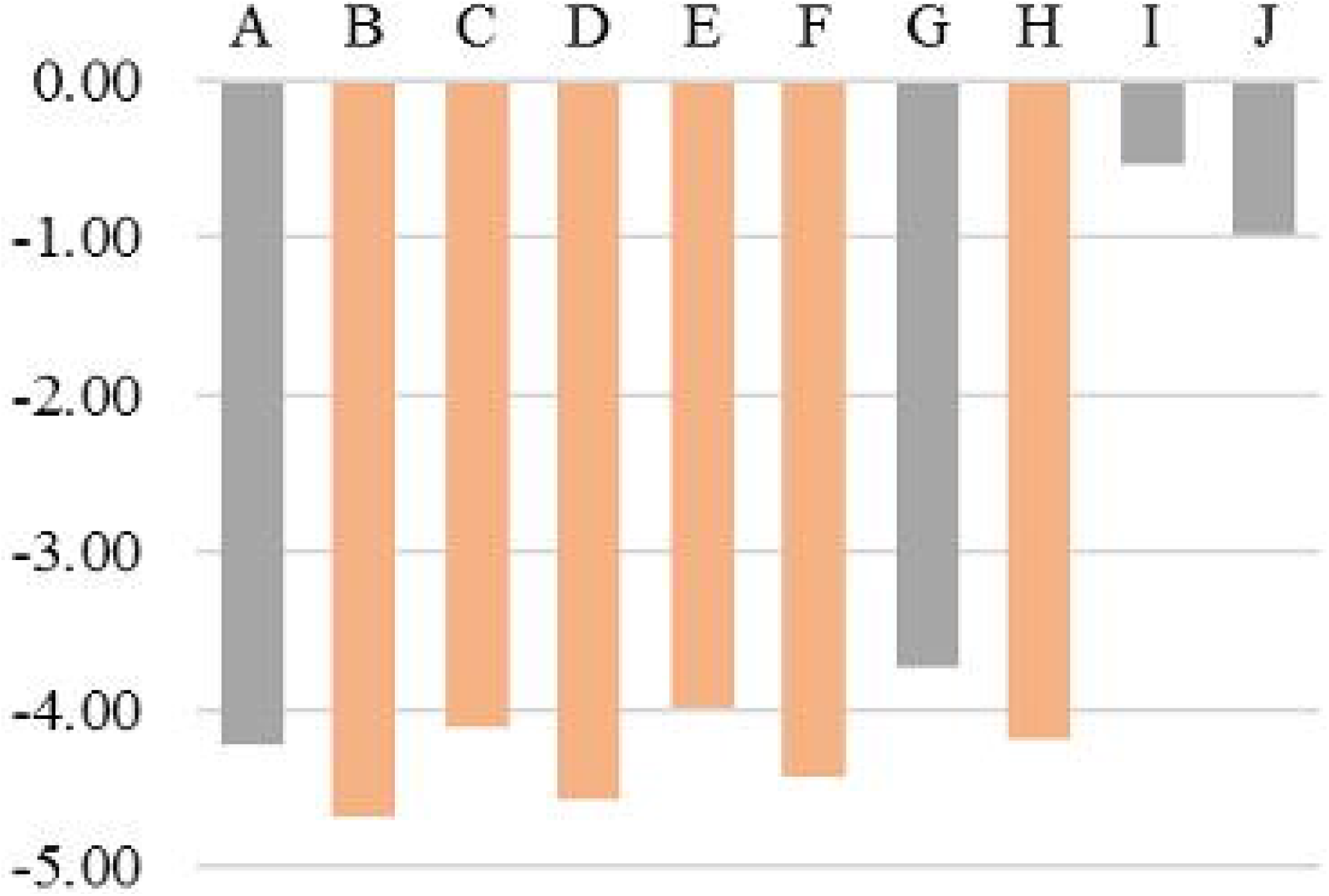
Bar chart of mean difference comparison between non-addiction and addiction group. *statistically significant (p < 0.05)

Additionally, as all significant models contained DLPFC, we performed a sub analysis using only DLPFC of both groups, and found that the results were not significant, both without and with ACC (p-value of 0.071 and 0.054, respectively).

## Discussion

Model C as classic definition of PFC was able to significantly distinguish pornography addiction from non-addiction group, along with model B, D, E, F, and H. The significant model B but not A, and model H but not G, showed that ACC was an essential region to distinguish pornography addiction as these pairs only differed in ACC. Model C, which also differed to model D in ACC but was similarly significant, has smaller effect size[26] (mean difference of model C vs D = -4.12 vs -4.57). ACC was reported to be important in craving,[27,28] self-control, inhibition, emotion regulation, motivation, self-awareness, attention, learning and memory. Disruption in ACC may result in impulsivity, compulsivity, and shifted motivation/attention into corresponding addiction.[9,29,30] Considering the important roles of ACC in addiction, we suggest that ACC would be included in future models of PFC on pornography addiction studies.

Significance in model C, compared to the not-significant model A, showed that IFG pars opercularis and triangularis, commonly referred as Broca’s area, was not required to distinguish pornography addiction. Although model B had the largest effect size among all models, it was inconsistent with the non-significant model A (similar in Broca’s area, differed in ACC which has been discussed above as essential region). Furthermore, model E-F pairs were similarly significant compared to model C-D pairs, even without IFG pars orbitalis, suggesting that this region was not an essential region. Our finding was in contrast with another study which claimed the role of IFG as a whole in addiction.[9] Additionally, comparison between model C-D pairs and E-F pairs suggested that addition of IFG pars orbitalis may increase effect size. Further studies specifying this region are required to confirm our finding.

Meanwhile, model G, which was short of OFC compared to model C, was not statistically significant, suggesting importance of OFC in pornography addiction. Model H (with ACC) was similarly significant compared to model D, but with smaller effect size (mean difference of model H vs D = -4.18 vs -4.57). OFC was known to be related to craving or pleasure expectation,[27] as well as emotion regulation/suppression, motivation, self-awareness, learning and memory.[9] The most important PFC region in pornography addiction might be DLPFC (SFG and MFG combined). This was observed from the not-significant model I-J pairs, which were DLPFC-short of C-D pairs. DLPFC was known to have important role in self-control, motivation, attention, working memory, learning, and decision making.[9] DLPFC might fit the pornography addiction model in juveniles, however by itself it could not distinguish addiction from non-addiction groups, as observed from the sub analysis involving DLPFC only.

One limitation of this study was its cross-sectional design, which cannot investigate cause-and-effect relationship. Additionally, brains of juvenile subjects are still growing,[31] therefore might compensate underlying brain damage.[32] Further neuroscientific studies using the models described above, especially on longitudinal designs, are warranted to find the best definition of PFC suitable for pornography addiction studies.

## Conclusions

The most suitable definition of prefrontal cortex for pornography addiction study in juvenile should consist of orbitofrontal cortex, anterior cingulate, and especially dorsolateral PFC. Inferior frontal gyrus pars orbitalis was not necessary for this purpose, but may increase effect size if it is included. Further studies are required to confirm these findings.

## Data Availability

fMRI data used to support the findings of this study are included within the article

## Abbreviation Lists

PFC: prefrontal cortex
DLPFC: dorsolateral prefrontal cortex
IFG: inferior frontal gyrus
OFC: orbitofrontal cortex (OFC)
ACC: anterior cingulate cortex
BPF‰: Brain Parenchymal Fraction per mille

## Additional information

No additional information is available for this paper.

## Acknowledgements

The authors would like to thank Alexandra Chessa, Resti Siti Saleha, Kevin Widjaja, and Nia Soewardi for their contributions in this paper.

## Authors’ contribution

Conceptualization: PuP, REE, HE. Investigation: PuP, REE, HE, SEIS, NZA, DC. Methodology: PuP. Formal analysis: PeP, GFH. Resources: HE, SEIS, NZA, DC. Writing (draft preparation, review, and editing): PeP, GFH, PuP. All authors critically reviewed and approved the final version of the manuscript.

