## Supplementary material for "Prefrontal Cortex Definitions and Their Use in Distinguishing Pornography Addicted Juveniles": Table 1

**Table S1. Each of questionnaire's components' mean and concurrent validity, averse to the three-step-norm.** To confirm the screening result, the 92-item questionnaire was followed by interview as qualitative input and also questionnaire about children tendencies in sexual activities (Table S2), namely the three-step-norm. Its validity and reliability were demonstrated by confirmatory factor analysis (CFA > 1.96) and Cronbach's Alpha (0.903; CA > 0.7) values.

| No. | Components | Mean ± SD | K-S p-value | p-value* |
|---|---|---|---|---|
| 1 | Sexual activities tendencies | 11.20 ± 11.20 | 0.000 | 0.023** |
| 2 | Number of times, frequency, and duration spent | 14.60 ± 2.54 | 0.121 | 0.013** |
| 3 | Motivation to use pornography | 20.50 ± 4.68 | 0.002 | 0.009** |
| 4 | Problematic pornography use | 69.55 ± 10.34 | 0.000 | 0.001** |
| 5 | Total score | 79.85 ± 30.01 | 0.858 | 0.000** |

*Unpaired t-test as the parametric test and Mann-Whitney as the non-parametric test.

**Statistically significant ($p < 0.05$)
