## Supplementary material for "Prefrontal Cortex Definitions and Their Use in Distinguishing Pornography Addicted Juveniles": table 2

**Table S2. Questionnaire of children tendencies in sexual activities.** In a scale of 1 to 5, it ranges from never to very frequent with minimum score of 11 and maximum score of 55.

| No. | Components |
|---|---|
| 1 | Masturbation |
| 2 | Embracing |
| 3 | Holding hands |
| 4 | Spend time together alone |
| 5 | Kissing |
| 6 | Hugging |
| 7 | Laying down together |
| 8 | Partner’s hands touching you in your attire |
| 9 | Your hands touching your partner in their attire |
| 10 | Laying down without clothes |
| 11 | Having sex |
